# Protocol for Generation of a Patient-Reported Outcome Measure of Quality of Life in Heart Valve Disease: The VALVQ

**DOI:** 10.1101/2023.05.20.23290285

**Authors:** Ariel Pons, Gillian Whalley, Crispin Jenkinson, David Morley, Sean Coffey

## Abstract

**Background:** There is an increasing prevalence of people worldwide with heart valve diseases (HVD), especially rheumatic heart disease, aortic stenosis, and mitral regurgitation, as well as people with a previous valve repair or replacement. Treatment decisions for HVD can be complex, making quality of life an important factor, but no questionnaire to measure quality of life across the lifespan of HVD exists. In this article, we describe the protocol for the development of such a questionnaire.

**Methods and Results:** The project will occur over four phases. First, people with HVD, family members and clinical experts will be interviewed to generate a list of questions (‘items’) that comprehensively describe participants’ quality of life. In the second phase, this will be formatted into a questionnaire that is pilot tested for functionality. In the third phase, items will be selected according to item distributions, factor analysis and rotation, and item response theory using the Graded Response Model to generate a final questionnaire containing only the best-performing items, which will then be tested for validity.

Validity assessments will be repeated after final questionnaire administration in a new sample in the fourth phase.

**Conclusion:** The article gives a template for development of a patient report outcome measure (PROM) in the health sciences. It is expected that the final questionnaire, called the VALVQ, will allow clinical trials to more sensitively assess quality of life changes across the spectrum and lifespan in HVD.

## INTRODUCTION

Heart valve disease (HVD) consists of a variety of conditions where the valves within the heart lose their ability to correctly direct blood flow, usually due either to excessive leaking (regurgitation) or restriction to flow (stenosis). HVD contributes to considerable morbidity and mortality worldwide, with an increasing prevalence seen over the past few decades(1). There is a growing cohort of people who have previously had a valve repair or replacement (VRR), but their epidemiology is much less well studied.

Although any heart valve can have clinically significant disease, three conditions have the largest impact: rheumatic heart disease (RHD), aortic stenosis (AS), and degenerative mitral valve disease causing mitral regurgitation (MR).(1) Rheumatic heart disease commonly occurs in young and socioeconomically disadvantaged populations. It is due to a infection by Group A Streptococcus which triggers an immunologic reaction, causing regurgitation and/or stenosis predominantly of the mitral valve and less commonly regurgitation of the aortic valve.(2) Aortic stenosis typically occurs in the elderly due to calcification of the valve, but can occur in younger people with a congenital malformation. Degenerative MR can occur in both the young and the elderly due to a wide range of primary and secondary causes.(1)

Presentations of these valve diseases vary but symptoms classically involve shortness of breath on exertion (or, in late-stage disease, shortness of breath at rest), angina, and fatigue.(3) Left untreated, HVD gradually leads to pressure and volume overload of the left ventricle, which eventually causes potentially fatal heart failure.(4) The treatment for HVD is valve repair or replacement (VRR). There are a wide range of replacement/repair modalities, necessitating the importance of QOL as an outcome in HVD. However, currently QOL cannot be sensitively measured in HVD since there is no existing questionnaire designed to measure QOL in HVD; existing studies of QOL in HVD use questionnaires developed in patients with heart failure or angina,(5, 6) or ‘generic’ questionnaires designed to compare across different diseases.(7, 8) One questionnaire has recently been generated to measure QOL in AS across transcatheter replacement,(9) but an instrument to measure QOL throughout the lifespan of HVD and across broader forms is required. In this article, we describe the protocol for development of a questionnaire to measure QOL in HVD, which we call the Valve Quality of life (VALVQ).

## METHODS

Standard psychometric practice in the development of PROMs was used, and in particular the evaluation of the instrument as described in the document “Patient-Reported Outcome Measures: Use in Medical Product Development to Support Labeling Claims” produced by the Food and Drug Administration, was followed.(10) The COREQ Checklist was used to guide reporting of this process, and is reported in the appendix.(11)

### Questionnaire development

The fundamental process of robust questionnaire generation is to first generate a questionnaire where all factors reported by participants which reflect changes in the disease state of interest and could relate to the outcome of interest are included. Secondly, the questionnaire is then shortened into the minimum number of best-functioning items. Generating the initial saturated questionnaire allows the experience of those with the disease state to be recorded; researchers cannot know which factors are important and which are not to people living with the disease state, and so must not rely on assumption. Subsequently shortening the questionnaire is necessary to produce a practical questionnaire and for the questionnaire’s statistical performance.

Questionnaire development will be split into four phases (Figure 1: Overview of study design). Phase one consists of item generation, where individuals with HVD, their families/carers, clinical experts, and results of a literature review will be used to form an item list that can be expected to cover all significant indicators of QOL in a HVD cohort. Pilot testing for basic functionality forms phase two, while questionnaire testing and dropping items form phase three, with phase four being validation.

**Figure 1.**
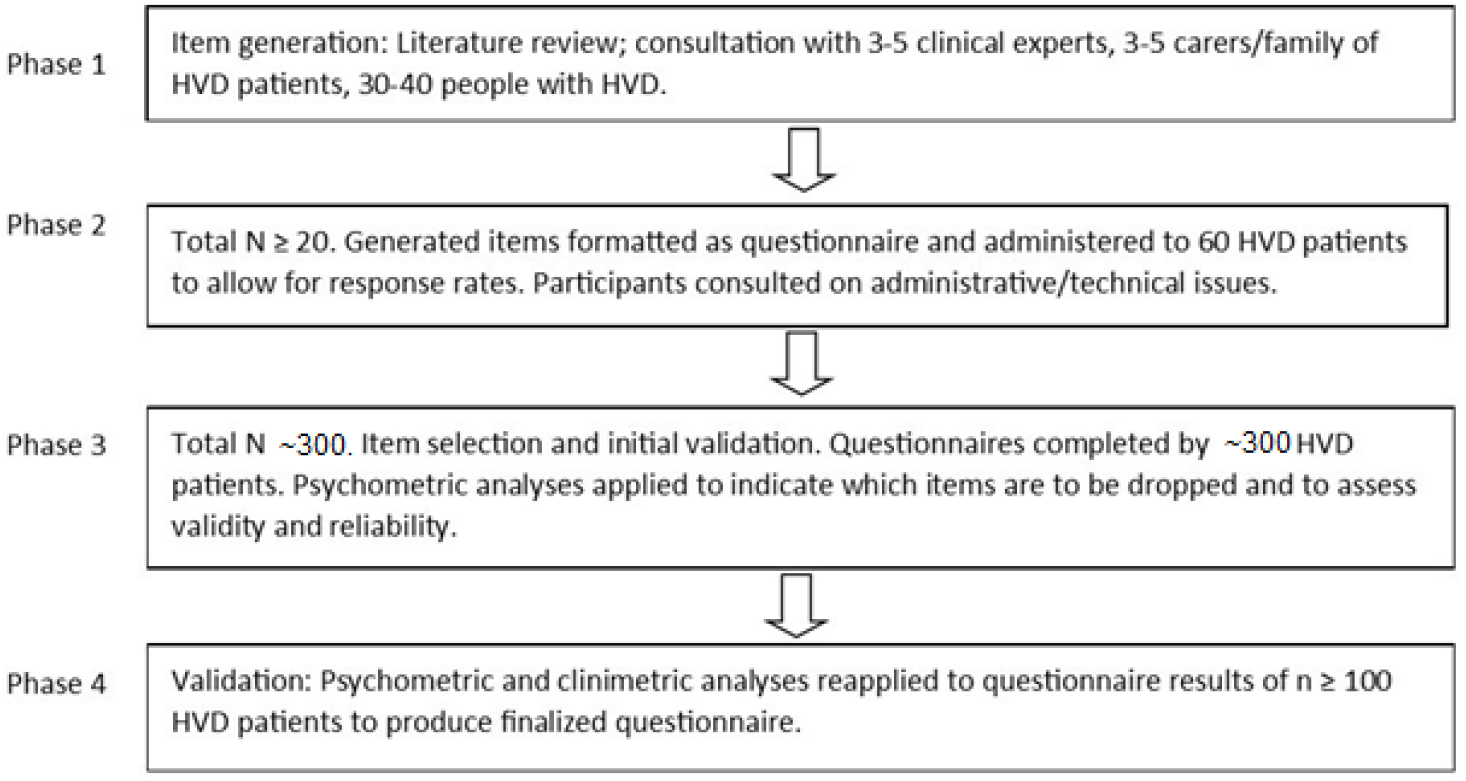
Overview of study design

#### Subject Inclusion and Exclusion

All participants will be aged over 18 years with a good knowledge of English and a diagnosis of one of the following forms of HVD: AS, MR, or RHD, or have undergone valve replacement/repair. The valve disease must be clinically significant which we define as moderate/severe AS or MR, any severity of clinically significant RHD, or any form of VRR. Participants will be excluded if they have a comorbidity more significant than their HVD, have recently been hospitalized, are pregnant (due to changes in heart function during pregnancy), are cognitively impaired, are deemed to be at risk of an impaired outcome from participating in this project, or do not consent to take part. All participants will be required to provide written informed consent. This project was approved by the New Zealand Health and Disabilities Ethics Committee (approval number 19/NTA/163).

#### Subject Screening and Enrolment

Individuals with HVD will be identified through echocardiographic databases of hospitals in New Zealand. Individuals deemed to meet the criteria will be contacted and sent an information sheet and consent form. Screening of consecutively screened participants will be performed to provide a representative sample of the VRR population undergoing echo.

### Questionnaire format

The questionnaire was formatted as semantic differential items with five-point scales, as this format better reflects QOL in this context. In contrast to the more common Likert scale, which has one statement with which respondents rate their agreement, a semantic differential presents paired bipolar opposing statements and the respondent answers by noting their position of relative agreement between the opposing statements. When measuring emotive concepts such as QOL, framing bias is a particular concern, but a Likert scale can only avoid framing bias by alternating the direction (i.e. positive vs negative wording) of its statements, which induces respondent error. The semantic differential format, with its opposing statements, has an inherently reduced risk of framing bias without an increase in error.(12)

Scales of QOL measures are typically either four or five-point scales. There are advantages and disadvantages to each: a four-point scale forces respondents to not tend towards the middle, but can cause participants to leave an increased number of questions unanswered.(13) A five-point scale does allow respondents to regress to the middle, but this reduces respondent burden. Since the semantic differential format chosen for this questionnaire already has a higher respondent burden, the questionnaire was created with a 5-point scale.

### Statistical methods

A sequential list of statistical methods is described in the results section – here we provide more detail on the methods used. This project will use factor analysis and item response theory (IRT) in its analyses. Factor analysis is the identification of underlying factors driving the variance in groups of items. Since we do not know how many factors there are in QOL in HVD – models of QOL in disease states vary widely in the number of factors (14, 15) – we used an exploratory factor analysis (EFA).(16) This reports the number of factors identified, in contrast to a confirmatory factor analysis in which a number of factors are input. EFA has two steps: first, the number of factors are identified in factor analysis, and then factor rotation is used to determine which items are grouped to which factor. Factor analysis and rotation is required before IRT, since IRT can only be applied within a single factor.(17)

Item response theory aims to explore the level of ‘latent trait’ – the variable being measured, e.g. QOL – in respondents. The model we chose provides detail on the function of individual items by generating discrimination and difficulty parameters for each item.’ Discrimination’ is an item’s ability to differentiate between respondents with different levels of the latent trait. ‘Difficulty’ is the level of latent trait required for a respondent to acquire higher scores on the item. Item parameters can be used not only for item selection when developing a questionnaire but also to investigate the functionality of the final questionnaire items. For example, parameters can assess whether responses are being driven by a factor other than the latent variable. This is done by assessing for differential item functioning (DIF).

Item parameters can also be used to generate scoring algorithms for the resulting questionnaire that can reflect respondents’ latent trait more accurately and make more use of complex data than classical test methods.(18) Item Response Theory, however, assumes univariance; that there is only one factor in the data. As such, IRT can only be performed after factor analysis and rotation, and the analysis must be performed separately within each factor. Discrimination parameters are also used here: while a high discrimination parameter indicates good item function, an excessively high discrimination parameter (four times than that of others)(17) indicates the violation of unidimensionality. IRT also assumes monotonicity, local independence, and invariance. Monotonicity is the assumption that, as a respondent’s level of latent trait increases, so does their probability of acquiring a higher questionnaire score; local independence is that, for each respondent, their answers to different items will be independent of other items, but reflect their latent trait; invariance is that item parameters are valid regardless of respondents’ scores.(19-21)

There is, at present, no strict consensus on the sample size calculation required for IRT. Some authors suggest a two-parameter model with a polytomous outcome on typically requires at least 400 participants to produce useful output,(22) but others have completed valid and useful studies with smaller sample sizes.(23, 24) Effective assessment of the latent variable requires a well-distributed dataset; a smaller dataset may likely be sufficient if the data are of good quality rather than skewed by biases or floor-ceiling effects. As such, we aimed for 300-400 participants with a variety of forms of HVD.

In this project, oblique factor rotation was selected as it is assumed that factors underlying QOL will correlate with each other. Our IRT model was chosen on the basis of which model best theoretically matched the structure of our data, rather than numerical fit.(21) We therefore selected the Graded Response Model (GRM), a logistic model designed for polytomous outcomes, as it is specifically designed for ordinal outcomes and especially scales that rate agreement,(35) and functions in practice at least as well as other polytomous models such as the partial credit model (PCM).(36) Moreover, while some consider the PCM to have more attractive statistical properties, especially for assessing latent variables in a population, our aim was to use IRT to select items – as the PCM does not generate a discrimination parameter, the GRM is superior for this task.(35) According to this selection by best theoretical structure, model fit was not calculated as it would not add value to our selection process.

To assess internal reliability, Cronbach’s Alpha was selected. This was selected as there is no consensus on the best reliability coefficient, and while the more computationally complex Macdonald’s Omega is being increasingly used, it has not been shown to be superior in practice to Cronbach’s Alpha.(37)

## RESULTS

### Phase One: Item Generation

In phase one, a comprehensive list of items assessing all aspects of QOL in HVD will be generated through three sequential steps: Systematic literature review of QOL in HVD; semi-structured interviews of people with different presentations of HVD to determine their opinions of indicators of their QOL; semi-structured interviews of clinical experts of HVD (n=3-5) and family/carers of people with HVD (n=3-5) to determine their opinions of factors that indicate patient QOL in HVD until saturation is reached.

Transcription of interviews will be done by the research team. The conclusions will be used to generate a comprehensive list of items that can be reasonably expected to reflect all significant indicators of QOL in HVD. This will be formatted into a self-reported questionnaire which will be computerized and designed to be completed on a smartphone or other electronic device. A printed paper version will also be made available.

### Phase Two: Pilot Testing

In order to identify any administration issues with the questionnaire, 20 questionnaires will be returned from participants with different forms of HVD. The questionnaire will also include a feedback form asking participants to identify any items they found difficult, unclear, insensitive, or irrelevant, and if there are any indicators of their QOL that have been missed. Items deemed clearly unsuitable in this phase may be dropped from the questionnaire, but the threshold to do so will be high as dropping items is to be done primarily in phase three.

### Phase Three: Initial item Selection

Item selection will involve a sample of approximately 300-400 people with different forms of HVD. Participants will also be asked to complete concurrent measures of QOL (the SF-20 and KCCQ) for reliability and validity assessments.(25)

The following analyses will be performed sequentially to indicate which items are to be dropped. Items may be retained despite the results of these tests if they are important contributors to the descriptive capacity of the questionnaire according to the results of phase one.

- The standard deviation (SD) for each item will be calculated, and items with a very low SD may be dropped (‘very low’ SD being determined by a distribution plot).
- Floor and ceiling effects will be described to assess item performance as well as whether the 1-5 range item scale is suitable.
- Factor analysis and factor rotation will be used to identify questionnaire structure. Factor analysis will be exploratory, as the number of factors is unknown. Factor rotation will be oblique, as it is assumed that factors of QOL will have some correlation with each other rather than being completely independent, and varimax, since this is the most commonly used method.(26)

### Phase Three: Item response theory analysis

After factor analysis, IRT will be used to compare items within each factor as follows:

- Discrimination parameters will be used to individually assess items; the distribution of discrimination parameters will be plotted to define low parameter values, and items with low discrimination parameters will be removed. Items with extremely high discrimination parameters (at least 4 times that of other items) will also be removed.
- Difficulty parameters within factors will be plotted to select between redundant items. Many items of the questionnaire will be different wordings of the same concept, as it is not known which wording will be found more relatable by respondents. Selecting between these ‘redundant’ items will be done by plotting difficulty parameters to see where the parameters are very similar. If two items have a similar difficulty parameter, the loss of one will not impact the ability of the questionnaire to measure the latent variable; if one item is a redundant item as described above and its differently-worded partner has a different difficulty parameter, then the item that has a similar difficulty parameter to any other item will be removed.

Information plots will be generated for each item. These indicate at which level of the latent variable the item performs best – i.e. if it performs best at negative versus positive QOL, or at extremes of QOL versus around the median.(27)

A scoring algorithm will be generated by weighted sum scoring; that is, the difficulty parameter for each level of response to an item will be multiplied by the discrimination parameter for the item as a whole, and then scaled to produce a score of 0-100.(28)

### Phase Three: Validation

Validity and reliability will be assessed once the final items have been selected.

- Differential item functioning will be used to assess the effect of gender and HVD status (native disease vs. replaced/repaired valve)
- Test-retest reliability will be determined longitudinally using a second administration of the questionnaire 2-3 weeks later, using only participants whose QOL according to the concurrent QOL measures has not significantly changed.
- Internal reliability will be calculated cross-sectionally with Cronbach’s Alpha.
- Local independence will be assessed by a correlation matrix of items’ residual covariance.
- Concurrent criterion validity will be assessed cross-sectionally by comparison of this questionnaire with SF-20 and KCCQ scores.
- At each administration of the questionnaire, feasibility will be assessed by response and completion rates. A codebook of ambiguous, unclear, incomplete, or missing answers will be compiled to identify any items that may need alteration.
- 20 randomly selected participants and 3-5 clinical experts will be interviewed to determine their opinions on the questionnaire.
- Linguistic validity will be assessed through concept elaboration documents and translatability assessment. Concept elaboration documents deconstruct items into the simplest possible sentences, which are then assessed by the research team as to whether the interpretation was as intended. Translatability assessments examine whether the wording of items is free of culturally-specific terms and could be easily translated into another language (rather than the separate process of actually translating a questionnaire.)(29)

## DISCUSSION

In this paper we describe an approach to developing a QOL measure that can be used across the spectrum and natural history of HVD. This approach can be used for development of other QOL measures. We initially focused on patients with AS, MR, RHD, and VRR, but envisage that the VALVQ can be used in other valve disease populations after appropriate testing.

While VRR reduces mortality in HVD, quality of life (QOL) is still an important outcome in HVD. Firstly, VRR has burdens of its own: replacement valves can be tissue valves, which have a limited life span and may need reoperation, or metallic valves, which last longer but require permanent anticoagulation.(30, 31) Secondly, there are many treatment options for HVD; intervention can be before or after overt symptoms develop, and can be performed with either surgical or transcatheter approaches, with a wide range of manufactured replacement valves available. QOL becomes an important guiding factor when there is no clear mortality advantage to a particular modality, and the improvements in expertise and quality of VRR ensures that most cases have multiple options which would be suitable, making QOL consideration an imperative. The literature reflects this, with an increasing number of studies assessing QOL after different valve treatment modalities.(32) Valve replacement also causes its own symptom burdens, such as the demand for long-term anticoagulation and blood tests when non-tissue valves are used, painful injections as part of RHD treatment, and recovery from surgical incisions. Finally, symptom burden is often not totally reduced after VRR,(33, 34) and clinical sequalae of living with HVD means that patients are often left with improved but still present heart failure symptoms.

## Limitations

Phase one, in which the QOL of people with HVD is elicited, only uses interviews and does not use focus groups. The addition of focus groups would have allowed the researchers to elicit data that they could not do themselves since they, unlike focus group participants, do not share the lived experience of HVD.(38) However, phase one of this project occurred during nationwide lockdowns for COVID-19, which made focus groups unfeasible due to many participants being unfamiliar with online group meetings. Secondly, HVD often occurs in people who are elderly who often have multiple comorbidities, many of which result in similar limitations to QOL as HVD. This makes assessment of QOL due purely to HVD difficult. Equally, there is wide spectrum of severity of HVD, from asymptomatic mild disease to end-stage disease, making measurement of the QOL underlying questionnaire responses difficult at the extremes of QOL. However, clinical decision-making at the extremes has a lesser requirement for quantification of QOL; treatment options have a lesser requirement of guidance by QOL. Assessment of the effect of clinical therapies on QOL is of more importance at intermediate levels of HVD.

As noted above, there is no consensus on sample sizes, and so this paper cannot advise future researchers on selecting a sample size.

## CONCLUSION

With the approach described here, we are confident that we can develop a QOL instrument for use in patients with HVD, both for individual patient care and for research. In particular, given the limitations of medical therapies in HVD, the VALVQ should allow more accurate measurement of participant-reported change in clinical trials.

## Data Availability

All data produced in the present study are available upon reasonable request to the authors

## ACKLNOWLEGEMENTS

This study was funded by the Department of Medicine, University of Otago, New Zealand.

## STATEMENTS AND DECLARATIONS

This study was funded by the Department of Medicine, University of Otago, New Zealand. Each author contributed significantly to the development of this protocol. Ethics was granted by the New Zealand Health and Disability Ethics Committee (Ethics reference 19/NTA/163). All participants gave informed consent to participate in this study, and those who participated gave consent to publish. No authors of this study had competing interests. This project was funded by the Department of Medicine, University of Otago.

